# Epidemic Landscape and Forecasting of SARS-CoV-2 in India

**DOI:** 10.1101/2020.04.14.20065151

**Authors:** Aravind Lathika Rajendrakumar, Anand Thakarakkattil Narayanan Nair, Charvi Nangia, Prabhal Kumar Chourasia, Mehul Kumar Chourasia, Syed Mohammed Ghouse, Anu Sasidharan Nair, Arun B Nair, Shaffi Fazaludeen Koya

## Abstract

**BACKGROUND:** India was one of the countries to institute strict measures for SARS-CoV-2 control in early phase. Since, then, the epidemic growth trajectory was slow before registering an explosion of cases due to local cluster transmissions.

**METHODS:** We estimated growth rate and doubling time of SARS-CoV-2 for India and high burden states using crowd sourced time series data. Further, we also estimated Basic Reproductive Number (R0) and time dependent reproductive number (Rt) using serial intervals from the data. We compared the R0 estimated from five different methods and R0 from SB was further used in analysis. We modified standard SIR models to SIRD model to accommodate deaths using R0 with the Sequential Bayesian method (SBM) for simulation in SIRD models.

**RESULTS:** On an average, 2.8 individuals were infected by an index case. The mean serial interval was 3.9 days. The R0 estimated from different methods ranged from 1.43 to 1.85. The mean time to recovery was 14 ± 5.3 days. Daily epidemic growth rate of India was 0.16 [95%CI; 0.14, 0.17] with a doubling time of 4.30 days [95%CI; 3.96, 4.70]. From the SIRD model, it can be deduced that the peak of SARS-CoV-2 in India will be around mid-July to early August 2020 with around 12.5% of population likely to be infected at the peak time.

**CONCLUSIONS:** The pattern of spread of SARS-CoV-2 in India is suggestive of community transmission. There is a need to increase fund for infectious disease research and epidemiologic studies. All the current gains may be reversed rapidly if air travel and social mixing resumes rapidly. For the time being, these must be resumed only in a phased manner, and should be back to normal levels only after we are prepared to deal with the disease with efficient tools like vaccine or a medicine.

## Background

SARS-CoV-2 originated in Hubei province of China and quickly spread to several countries including Japan and South Korea.^1^ The source of the infection was later traced back to wet market in Wuhan, capital of Hubei.^2^ A phylogenetic analysis of virus in Italy confirmed likely import of disease from China.^3^ Rapid travelling modes such as air transport further accelerated the epidemic by introducing new cases in to more countries.^4^ As of April 13 2020 there were 1,922,891 reported cases of SARS-CoV-2 globally along with a huge death toll almost touching 120,000.^5^ There is increasing evidence that the virus may have jumped organisms to reach humans through pangolins or similar animals acting as intermediates.^6^

Emerging and re-emerging infections of this scale disrupt health system functioning and cause massive losses to economy. Mathematical modelling has been used widely to understand the spread of disease in populations.^7^ One of the aims of the modelling is to estimate parameters that are critical to the spread of diseases such as basic reproductive number or R0 and incubation period.^8^ This is quite useful in the scenario of early spread of disease to plan future strategies.

India was one of the countries to institute strict measures for SARS-CoV-2 control in early phase.^9^ The first case of SARS-CoV-2 in India was reported on January 30 2020 from a student airlifted from Wuhan and at the same time another 800 suspected individuals kept under observation.^10^ Since then, the epidemic growth trajectory was slow before registering an explosion of cases due to local cluster transmissions. In this paper, we describe the current trend of SARS-CoV-2 transmission in in India and estimate basic parameters such as basic reproductive number R0 and time reproductive number Rt, doubling time and future trend for India from real world datasets. Our findings will help to understand the effectiveness of government response and recalibrate suppression strategies for the epidemic.

## Methodology

We used crowd sourced time series data available from the internet to estimate country specific parameters for the epidemic.^11^ As of now, this is the best available database for information on SARS-CoV-2 in India in public domain. The most reliable information in this database is regarding the daily reported incidence of COVID cases. Also, it contains aggregated information on total confirmed cases, total death and total recovery. We did not use variables with limited information. The data was scraped on to R software on 12^th^ April 2020, cleaned and reshaped for the current analysis.^12^ Package ggplot2 was mainly used to create figures along with features from base R.^13^ We defined serial interval as the time difference in diagnosis of SARS-CoV-2 in infectee and infector.^14^ We assumed serial interval and generation time to be same. Growth rate was computed from this incidence curve by fitting a log-linear model ^15^ and basic reproduction number (R0) was obtained from previously calculated serial interval. We also calculated time dependent reproductive number (Rt) to show the change in infectivity over time. State-wise growth was measured using the slope from a linear model.To estimate an R0 for projection purposes, in addition to log linear, we also estimated R0 from other methods such as Exponential growth (EG), Maximum likelihood estimation (ML) and Sequential Bayesian Method (SB) and Estimation of time dependent reproduction numbers (TD).^16^ We selected Sequential Bayesian Method (SB) to account for stochasticity in incidence curves as the priors change with time and are drawn from posterior distribution from time dependent informative priors.^16^

Conceptually, susceptible individuals become infectious (i.e., move from the susceptible compartment to the infectious compartment), and then ultimately recover from the infection (i.e., move from the infectious compartment to the recovered compartment). The rates at which they move from one compartment to another depend on the proportion of the population in each of these compartments, as well as the transmission and recovery rates associated with the disease. We modified standard SIR models to accommodate an additional compartment for deaths by assuming transition probabilities, the susceptible-infectious-recovered/Death (SIRD) model. Our model can be represented by the following ordinary differential equation

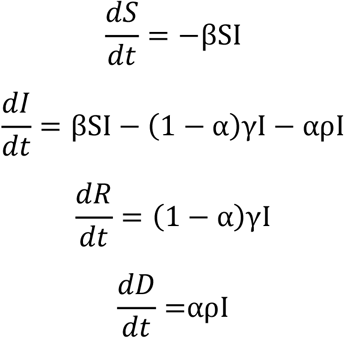

where β is the disease transmission rate, γ is the recovery rate, α is the case fatality rate and ρ rate at which death occurs. β is the product of contact rate and transmission probability. As these parameters could not be estimated directly from the data, we back calculated β from R0 which was computed previously using Sequential Bayesian method. γ was calculated from infectious period - (1/ infectious period). Case fatality rate (α) can be defined as deaths out of confirmed cases. ρ was estimated as inverse of time to death. We did not have ρ from our data and hence used estimated range of 2-8 weeks from the WHO report.^17^ We used 5 weeks as the value of ρ for our study. The values of the parameters are as follows: R0=1.85, b=0.13, γ =0.07, α=0.035 and ρ=0.028.

## Ethics Approval

We used anonymised data available in the public domain for this analysis.

## Results

The first case of SARS-CoV-2 was reported in Kerala state. Since then as of 12 April 2020, there are 9212 confirmed cases, 248 deaths and 1226 recovered in India from SARS-CoV-2. Maharashtra report the highest number of cases (1982) and north eastern states have the lowest reported cases. The trend of the epidemic is shown in Figure 1.

**Fig 1:**
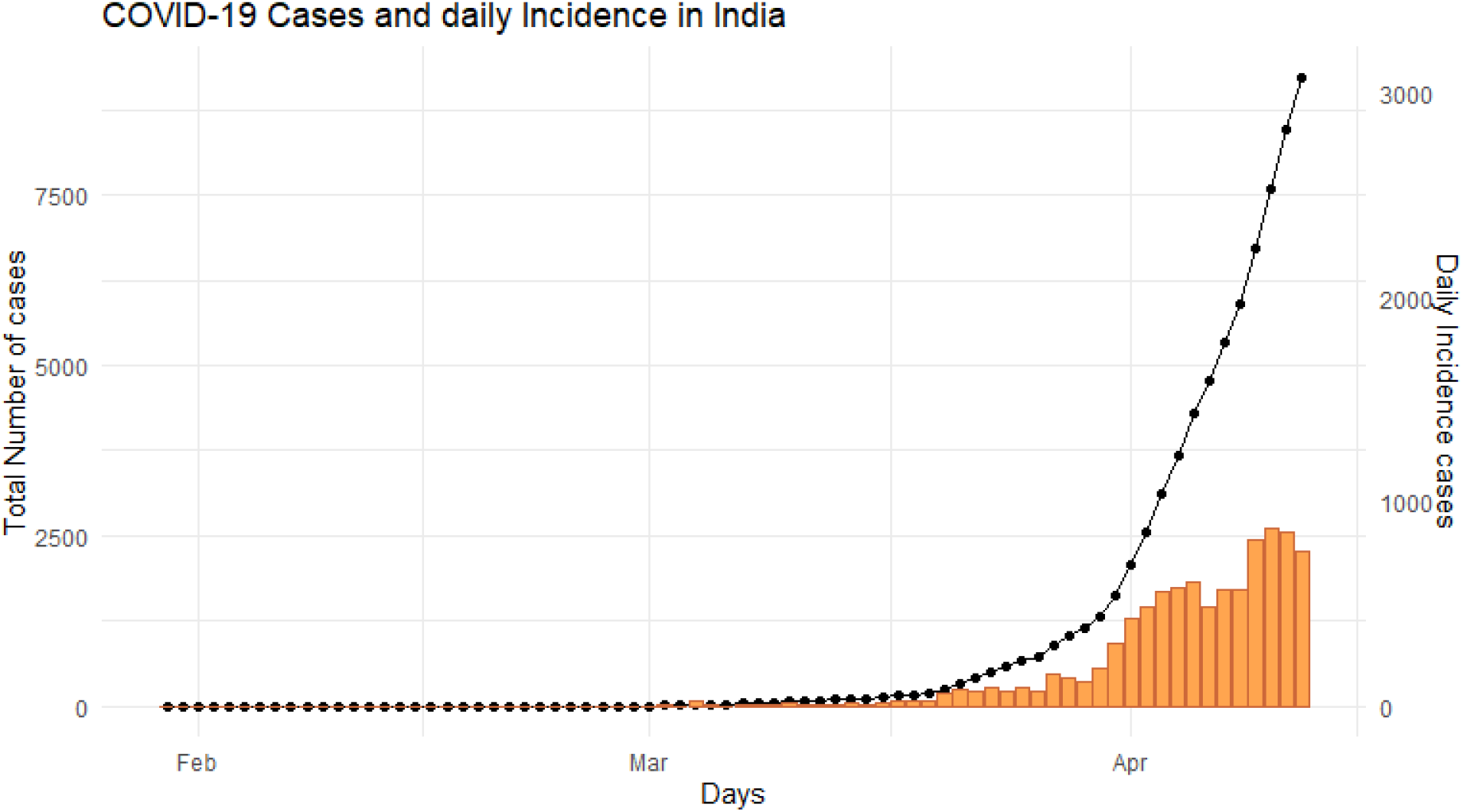
Daily and cumulative incidence of SARS-COV-2 cases in India

Daily growth rate of India fitted from a log incidence over time is 0.16 [95%CI; 0.14, 0.17]. The doubling time for the epidemic is 4.30 days [95%CI; 3.96, 4.70]. The growth rate and doubling time for the 5 states with highest burden are given in Table 1 and the rates for the states with available data are given in Supplementary Figure 3. Kerala, which had the highest number of cases have effectively contained the outbreak by prolonging the doubling time, 19.48 days [95% CI;10.35,164.60] and the present growth rate is 0.03 [95%CI; 0.004, 0.066].

**Table 1:** Growth rate and doubling curve from log-linear fit for 5 states with highest burden of SARS COV-2 in India

| State | Growth rate (95% CI) | Doubling time in days (95% CI) |
| --- | --- | --- |
| Tamil Nadu | 0.173 (0.13,0.20) | 4.00 (3.32,5.03) |
| Delhi | 0.145 (0.12,0.17) | 4.78 (4.05,5.83) |
| Maharashtra | 0.144 (0.12, 0.16) | 4.81 (4.23, 5.58) |
| Rajasthan | 0.131 (0.11,0.15) | 5.27(4.61,6.17) |
| Gujarat | 0.119(0.07,0.17) | 5.77(4.13,9.59) |

The nature of the epidemic is changing as the transmission is now more prominent with in local communities as opposed to imported cases from individuals with travel history from China, Middle East and Europe. The type of transmission for most cases were not available. However, those individuals whose source of infection is yet to be determined can be assumed to be of local origin as travel history of SARS COV-2 patients is well tracked and documented in India (Supplementary Figure 2).

There were 145 index cases who were identified to have transmitted the disease. On an average, 2.8 individuals were infected by an index case and the range varied from 1-20 individuals (Figure2). We had information of 413 pairs of primary and secondary cases to generate serial intervals for computing Basic Reproductive Number or R0. Serial interval ranged from 0-19 days as the primary and secondary cases reported to the health facility on the same date. The mean of serial interval was 3.9 days with a standard deviation of 2.85 computed from pair of 324 pairs with non-zero values

**Figure 2.**
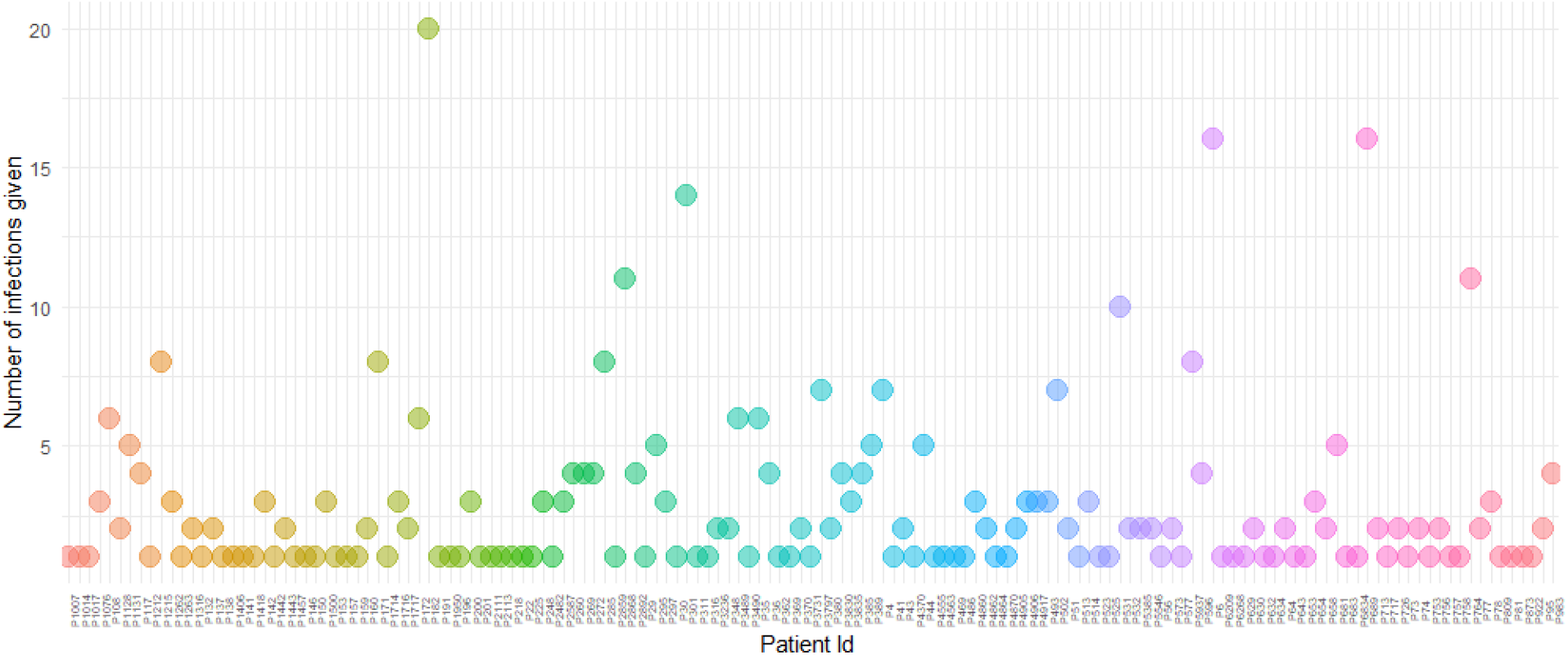
Number of Secondary cases resulting from contact with each index case

The lowest estimated R0 was from log linear model 1.47 [95% CI, 1.43, 1.51] and highest from time dependent model 1.89 [95% CI, 1.64-2.15] (Figure 3). The time varying reproductive number (Rt) and SI distribution curve across the analysis time is provided in Supplementary Figure 1. The recovery time was available only for 133 individuals. The mean time of recovery was 14 days (SD 5.3) ranging from 5 to 25 days.

**Figure 3:**
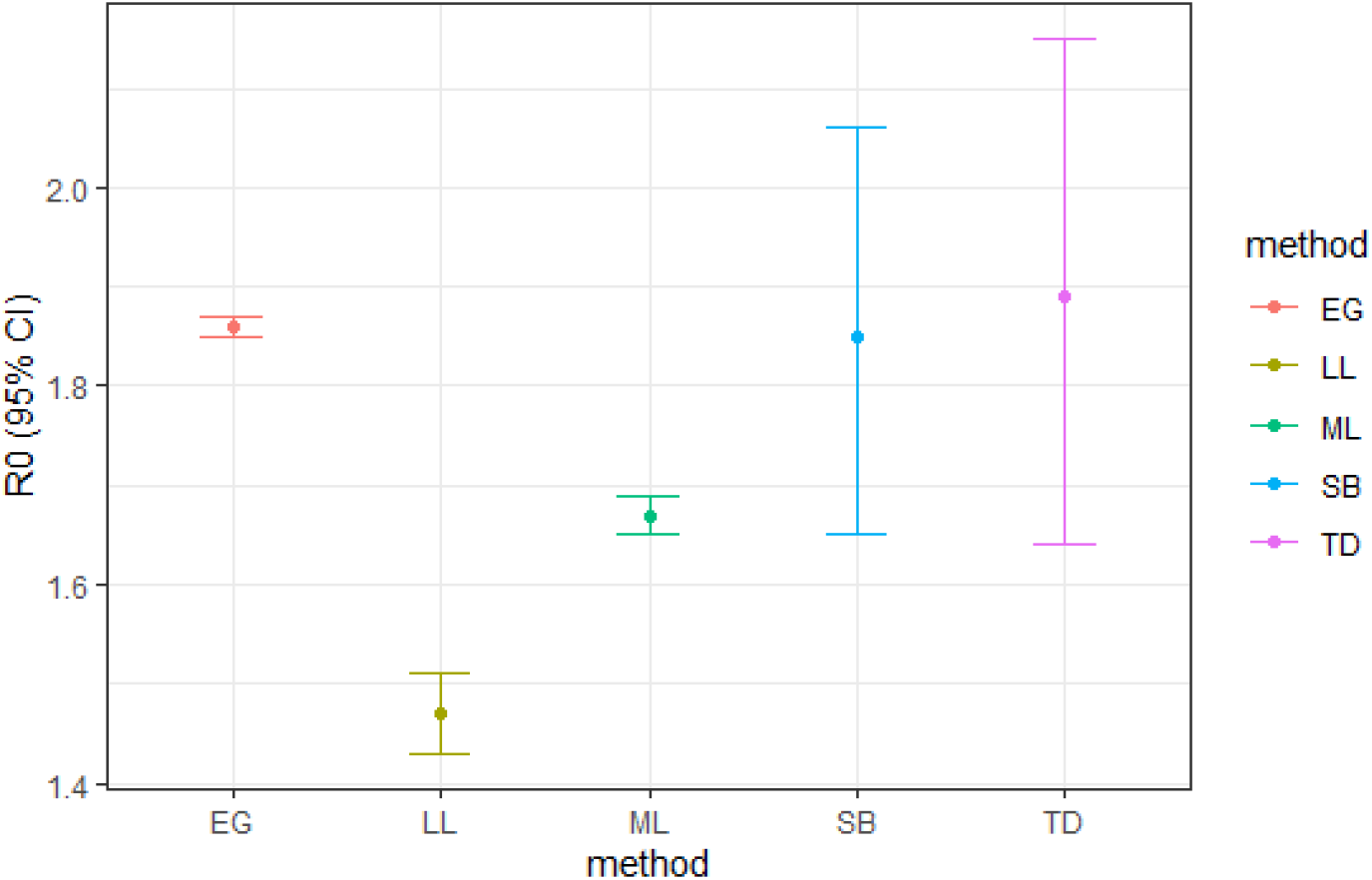
Basic reproductive number (R0) calculated from different estimation methods

From the compartmental models it can be deduced that the peak of SARS-CoV-2 in India will be around mid-July to early August 2020. Around 12.5% of the susceptible individuals are likely to be infected during the peak. The R0 estimated from different methods ranged from 1.43 in log-linear method to 1.85 in exponential growth methods. Since the estimates were similar between exponential, Time dependent and Sequential Bayesian methods we went forward with the Sequential Bayesian to create the SIRD model (Figure 4).

**Figure 4.**
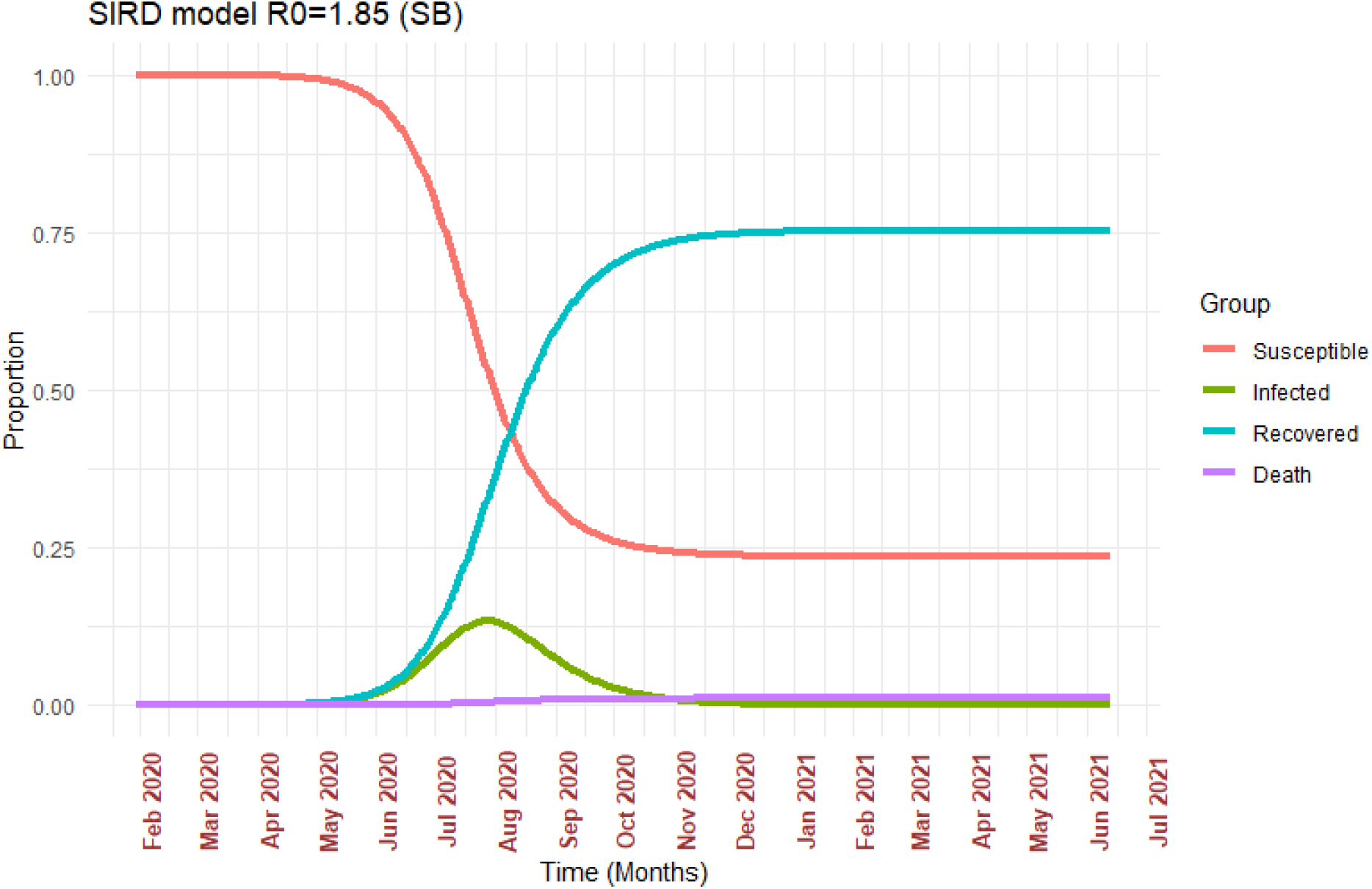
SIRD models based on R0 derived from Sequential Bayesian Method (SBM)

## Discussion

We estimated several parameters of SARS-CoV-2 infection in India, from crowd sourced data and used minimal set of assumptions in our forecasts. Models are useful in identifying the transmission dynamics if parameter inputs are based on real world data in early phase of epidemic. Social distancing and travel ban to reduce community transmission seem to have impact on infection transmission.

There is an urgent need to estimate setting specific parameters for future planning. For instance, we did not have information on death rate and recovery rate and used estimates from published studies in the SIRD model. We also could not compute incubation period as the database contained information only on linked cases and their admissions. Studies have shown that this could be approximately between 5-8 days.^18,19^ Also, it is difficult to separate the serial interval time from generation time overlaps as the information of cases are only available from reporting to the health authorities. Studies have suggested that SARS-Cov-2 can spread before the symptomatic phase which precludes full understanding of the transmission pattern and underlying characteristics.

The R0 estimated across many studies may not reflect actual transmission potential of virus as these were measured in dynamic cohorts with some sort of mitigation measures. Estimates from studies among closed population such as in Diamond Princess and Wuhan may be more reliable in this regard.^20^ Riou et al. estimated a reliable range for R0 which was between 1.4 and 3.8 by simulating Wuhan epidemic incidence trajectories in a cluster environment.^21^ We estimated the range of R0 in India to be between 1.42 and 1.84 by comparing estimates from 5 different methods. It can be assumed that the real Ro in India is close to 1.85. The upper limit of uncertainty of R0 in our analysis exceeds 2 which may be indicative of spread of infection at local level and is comparable to European scenario.^22^ This is much lower than R0 in most countries and may be indicative of the effectiveness of measures taken at policy level such as lock down, cancellation of flights, social distancing and sealing of identified hotspots.

India has done well to contain population level spread to a great extent and to keep low mortality rates. India has one of the lowest attack rate of 0.47 at this stage of the epidemic which is several times lower than developed countries.^23^ However, the epidemic is on the rise in India and creates new challenges such as optimal use of resources including swabs and testing kits besides identifying the high-risk groups. A recent study by Prem et al suggests that mitigation measures may be helpful only in delaying the infection from reaching its peak.^24^ Thus, it implies that the measures would likely prevent immediate flooding of hospitals preventing people from obtaining adequate care. The epidemic is yet to reach its peak in India which we expect to be around mid-July and early August. Our models suggest that 12.5% of the susceptible individuals are likely to be infected during the peak. This means that there is still around 2-3 months to prepare for the worst phase of the epidemic.

Control measures of SARS-CoV-2 are less effective in populations with high density.^20^ India has a huge population and potential of SARS-CoV-2 for sustained transmission is well established. ^21,25^ We relied solely on estimates from compartmental model which generates estimates from the infection parameters than depending on forecasts based on trend. Our findings are based on simulations and further measures at policy level can alter the trajectory of the disease. Lipsitch et al recommends a strategy combining epidemiological studies, testing and lab studies as more effective strategy in generating evidence for suppressing the epidemic.^26^

Our analysis has certain limitations. Firstly, there may be underreporting of cases and lack of validated data for research purposes available to the public.However, we compared the incidence from our dataset to the aggregated numbers elsewhere like European Disease Prevention and Control ^23^and Worldometer^5^ and the figures were similar. Another issue in estimating the parameters is the presence of super spreaders and asymptomatic cases. The estimated proportion of asymptomatic cases can be as high as 10% in the population.^27^ The virus can be spread around during asymptomatic stage and delays from onset to reporting or treatment can be as high as 11 days.^28^ Nonetheless, pooling estimates across several studies will aid in computing values that are closer to the true estimates.

In conclusion, our study provides information on India specific estimates of SARS-CoV-2 transmission parameters using real world data for the first time and shows that measures taken till date have been effective in reducing the spread of disease. However, the rising incidence and pattern of spread is suggestive of community transmission and is likely to increase cases in the future. The availability of individual level data is critical to assess the effectiveness of ongoing measures and plan future strategies. There is a need to increasingly fund infectious disease research and epidemiologic studies and make that data available in the public domain. Future studies could focus on studying genetic variations in asymptomatic individuals and those index cases with higher disease transmission rates. There needs to be a renewed thinking of health systems particularly regarding emergency preparedness and optimal utilization of scarce resources. Epidemiologists need to be considered in decision making and developing disease control strategies. Furthermore, there needs to be concerted action at global level to contain the virus including joint research and developing vaccines and drugs. All the current gains may be reversed rapidly if air travel and social mixing resumes rapidly. For the time being, these must be resumed only in a phased manner, and should be back to normal levels only after we are prepared to deal with the disease with efficient tools like vaccine or a medicine.

## Data Availability

Data is available in the public domain

## Acknowledgement

We acknowledge Dr Mike Lonergan, University of Dundee, UK for reviewing and providing critical inputs for the manuscripts. We thank the team managing COVID19 open data source which was used for this analysis.

---

**Supplementary Figure 1.**
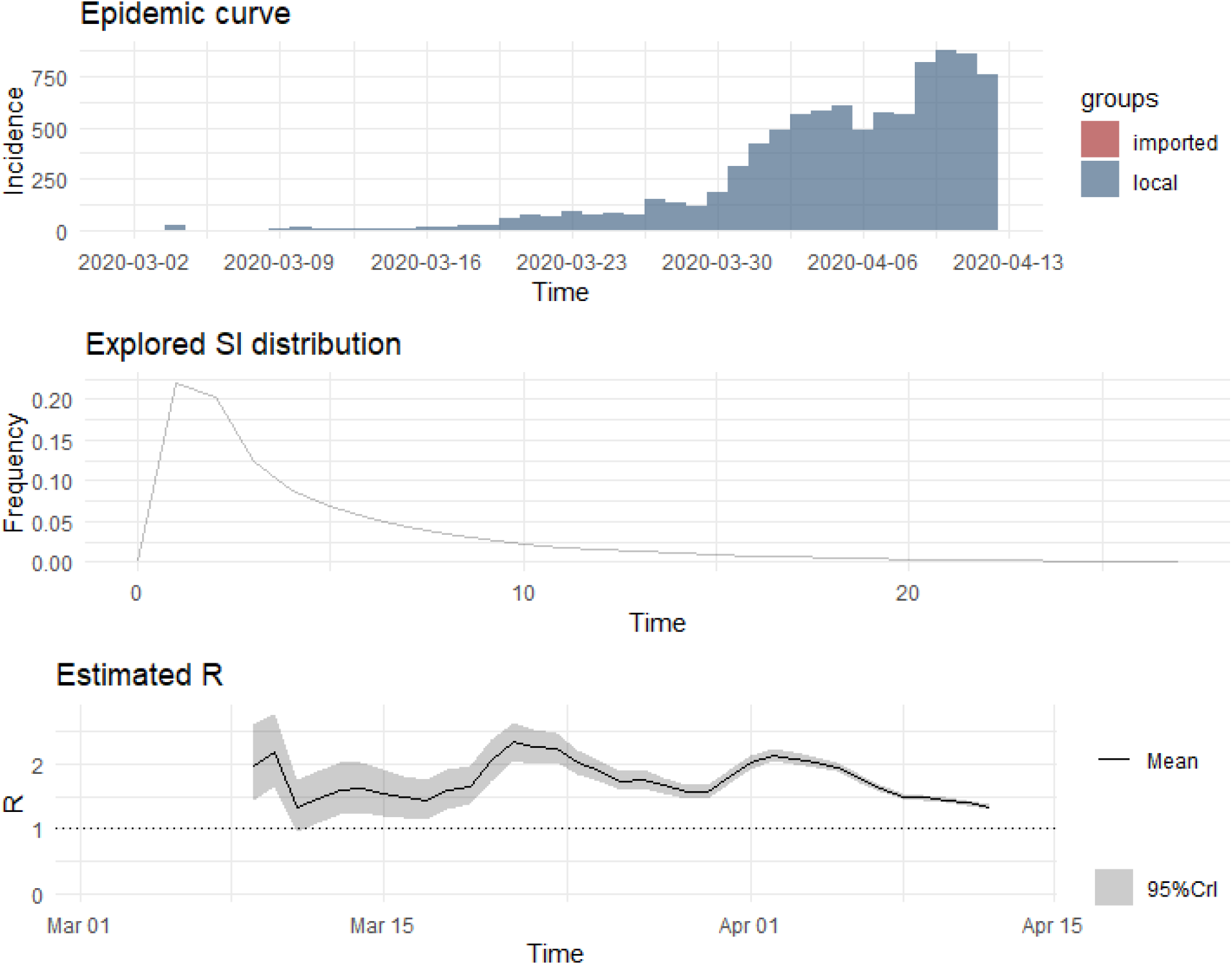
Epidemic curve, SI distribution and time varying reproductive number (Rt)

**Supplementary Figure 2:**
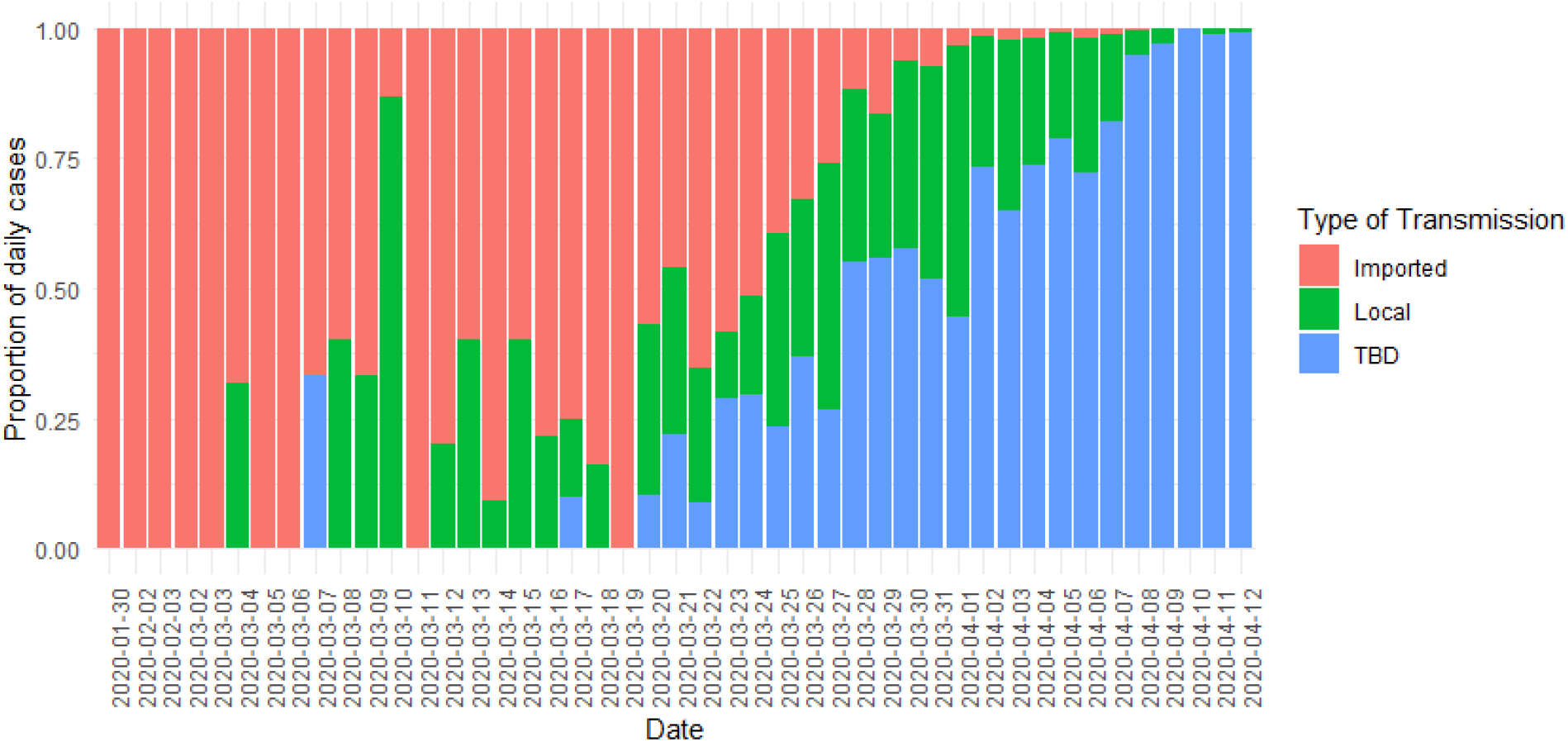
*Daily incidence and type of* SARS COV-2 *transmission* in India

**Supplementary Figure 3:**
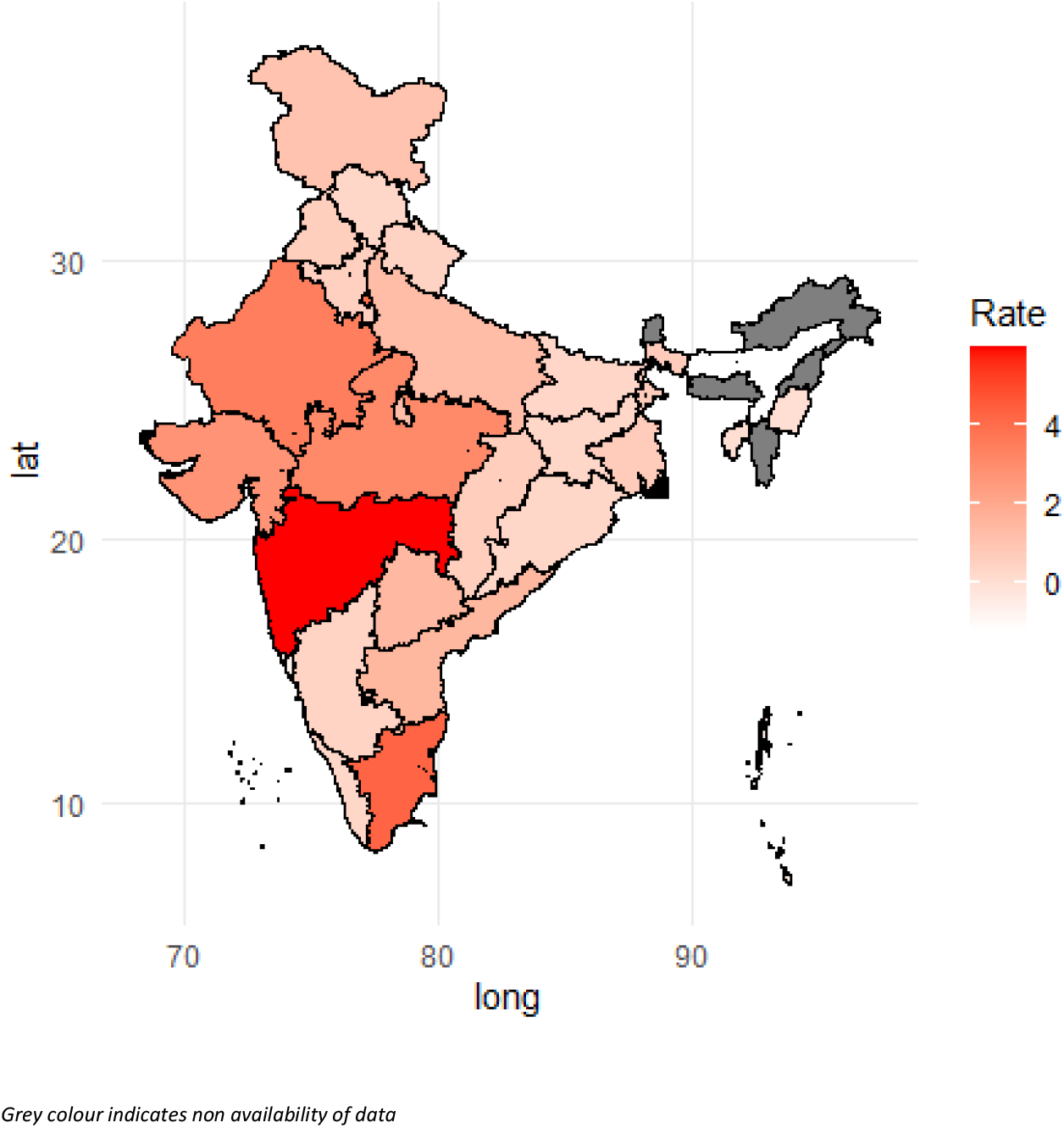
Growth rate in each state in India fitted with linear model

